# Neighborhood Social Vulnerability and Interstage Weight Gain: Evaluating the Role of a Home Monitoring Program

**DOI:** 10.1101/2023.03.13.23287234

**Authors:** Rachel J. Shustak, Jing Huang, Vicky Tam, Alyson Stagg, Therese M. Giglia, Chitra Ravishankar, Laura Mercer-Rosa, James P. Guevara, Monique M. Gardner

**Affiliations:** Division of Cardiology, Department of Pediatrics, The Children’s Hospital of Philadelphia and Perelman School of Medicine, University of Pennsylvania, Philadelphia, Pennsylvania; Department of Biomedical and Health Informatics, Data Science and Biostatistics Unit, The Children’s Hospital of Philadelphia, Philadelphia, PA; Cartographic Modeling Lab, University of Pennsylvania, Philadelphia, PA, USA; Division of Cardiac Critical Care Medicine, The Children’s Hospital of Philadelphia and Department of Anesthesiology and Critical Care, Perelman School of Medicine at the University of Pennsylvania, Philadelphia, Pennsylvania; Division of General Pediatrics, Department of Pediatrics, The Children’s Hospital of Philadelphia, Perelman School of Medicine at the University of Pennsylvania, Philadelphia, Pennsylvania

**Keywords:** Hypoplastic left heart syndrome, health disparities, interstage period, growth, congenital heart disease

## Abstract

**Introduction:** Poor interstage (IS) weight gain is a risk factor for adverse outcomes in infants with hypoplastic left heart syndrome (HLHS). We sought to examine the association of neighborhood social vulnerability and IS weight gain and determine if this association is modified by enrollment in our institution’s Infant Single Ventricle Management and Monitoring Program (ISVMP).

**Methods:** We performed a retrospective single-center study of infants with HLHS before (2007-2010) and after (2011-2020) the introduction of the ISVMP. The primary outcome was IS weight gain, and the secondary outcome was IS growth failure. Multivariable linear and logistic regression models were used to examine the association between Social Vulnerability Index (SVI) and the outcomes. We introduced an interaction term into the models to test for effect modification by ISVMP.

**Results:** We evaluated 217 ISVMP infants and 111 pre-ISVMP historical controls. SVI was associated with IS growth failure (P = 0.001), however, enrollment in ISVMP strongly attenuated this association (P = 0.04). Pre-ISVMP, high and middle vulnerability infants gained 4 gm/day less and were significantly more likely to experience growth failure than low vulnerability infants (high vs. low: aOR 12.5; 95% CI 2.5-62.2; middle vs. low: aOR 7.8; 95% CI 2.0-31.2). After the introduction of the ISVMP, outcomes did not differ by SVI tertile. Middle and high SVI infants enrolled in ISVMP gained 4 gm/day and 2 gm/day more, respectively, than pre-ISVMP controls.

**Conclusion:** In infants with HLHS, high social vulnerability is a risk factor for poor IS weight gain. However, enrollment in ISVMP significantly reduces growth disparities.

## INTRODUCTION

Hypoplastic left heart syndrome (HLHS) is a complex congenital heart defect associated with significant morbidity and mortality. The interstage period, defined as the time from stage 1 Norwood palliation (S1P) hospitalization discharge to the time of stage 2 palliation (S2P) is a tenuous time for infants with HLHS. Fluctuations in systemic and pulmonary blood flow due to residual lesions or intercurrent illness can lead to decompensation and if undetected, death (1). In recent years, social determinants of health have emerged as important predictors of interstage mortality (2–4). Despite these findings, considerable knowledge gaps remain in the understanding of interstage health disparities. Over the past decade, home monitoring, a strategy aimed to detect physiologic variances that precede clinical deterioration, has been widely adopted and resulted in a significant reduction in interstage mortality (1,5–7). However, nearly all studies of socioeconomic status (SES) to date have evaluated patient cohorts that pre-date the era of home monitoring. Furthermore, analyses have focused on the outcome of interstage mortality. However, with morality rates now as low as 2-5%, we must focus our attention on interstage morbidities (6,8,9).

One important interstage morbidity is growth failure (10,11). In infants with HLHS, weight gain is an important indicator of well-being and impaired growth can be an early clinical sign of decompensation (12). Poor interstage weight gain has been associated with longer hospital length of stay at the time of S2P and worse early developmental outcomes, as well as one-year transplant-free survival following S1P (13–15). Identifying risk factors for growth failure may result in improved outcomes. To date, there has been no in-depth investigation into the impact of social determinants of health on interstage weight gain.

In this context, the objective of this study was to examine the impact of neighborhood social vulnerability on interstage weight gain and determine whether this association is modified by enrollment in a standardized home monitoring program. We hypothesize that high neighborhood social vulnerability is a risk factor for poor interstage weight gain and growth failure. However, interstage home monitoring may have the ability to attenuate disparities by improving parental education prior to discharge along with communication and access to care after discharge.

## METHODS

The study protocol was approved by the Institutional Review Board for the Protection of Human Subjects at the Children’s Hospital of Philadelphia IRB# 19-017193. Requirements for informed consent were waived. The data, analytic methods, and study materials will not be made available to other researchers for purposes of reproducing the results or replicating the procedure.

### Study Population

We conducted a quasi-experimental interrupted time series analysis of patients with HLHS and HLHS variants who underwent S1P from January 1, 2007, to December 31, 2020. Our center’s home monitoring program, the Infant Single Ventricle Management and Monitoring Program (ISVMP) was introduced in January 2011. The ISVMP cohort was comprised of all infants with HLHS who underwent S1P after December 1, 2010 and completed their interstage period prior to December 31, 2020. The historical control cohort was comprised of all infants with HLHS who underwent a S1P between January 1, 2007, and November 30, 2010 (last date before the introduction of the ISVMP). Patients who underwent S1P in anticipation of a biventricular repair, those who died during the neonatal hospitalization, those who remained inpatient during the interstage period, those who resided internationally, those who were discharged on palliative care, and those with a missing S1P discharge weight or date were excluded. Given the variability in home monitoring programs, patients enrolled in outside hospital programs were also excluded.

### Home Monitoring Program

All infants enrolled in the ISVMP were discharged home with a pulse oximeter and weighing scale. The additional components of the ISVMP included: (1) standardization of the neonatal discharge criteria and process; (2) standardization of parental education before neonatal discharge; (3) daily monitoring by parents, including daily oxygen saturation measurement, daily weight measurement, and feeding log; (4) home nursing visits documenting oxygen saturation, weight, mode of feeding, and formula type and amount; (5) weekly phone calls to parents by a dedicated ISVMP nurse practitioner; (6) involvement of a registered dietician; (7) biweekly pediatrician visits; (8) biweekly cardiology visits with focused echocardiograms; (9) weekly review of patients by a dedicated ISVMP team; and (10) scheduling of standard S2P at 3 to 4 months of age. Standard criteria for remaining inpatient interstage included a continued need for inotropic support, respiratory support, or sedative medications, and arrhythmias refractory to standard medical therapy. Other criteria were at the discretion of the ISVMP team. There was no dedicated ISVMP outpatient clinic, therefore, cardiology visits were scheduled with primary cardiologists within or outside our center. Prior to the introduction of the ISVMP, S1P discharge criteria were determined by individual inpatient providers and frequency of follow-up was at the discretion of primary cardiologists.

### Neighborhood Social Vulnerability Index

Neighborhood social vulnerability is defined as the resilience of a community when confronted by external stresses on human health (16,17). Neighborhood social vulnerability was measured with the Centers for Disease Control Social Vulnerability Index (SVI). While initially designed to aid local governments in identifying vulnerable communities during natural disasters, the SVI has recently been applied more broadly in the investigation of healthcare disparities and emerged as a significant predictor of health outcomes (18–22). The SVI is derived from summed percentile rankings of fifteen United States Census variables. Variables are grouped into four themes that represent subcategories of SVI: socioeconomic status, household composition, minority status and language, and housing type and transportation (Table 1). Each census tract receives a separate percentile ranking for each of the four themes as well as a composite ranking. The SVI takes on values between 0 and 1 with higher values representing greater vulnerability than lower values (16).

**Table 1:**
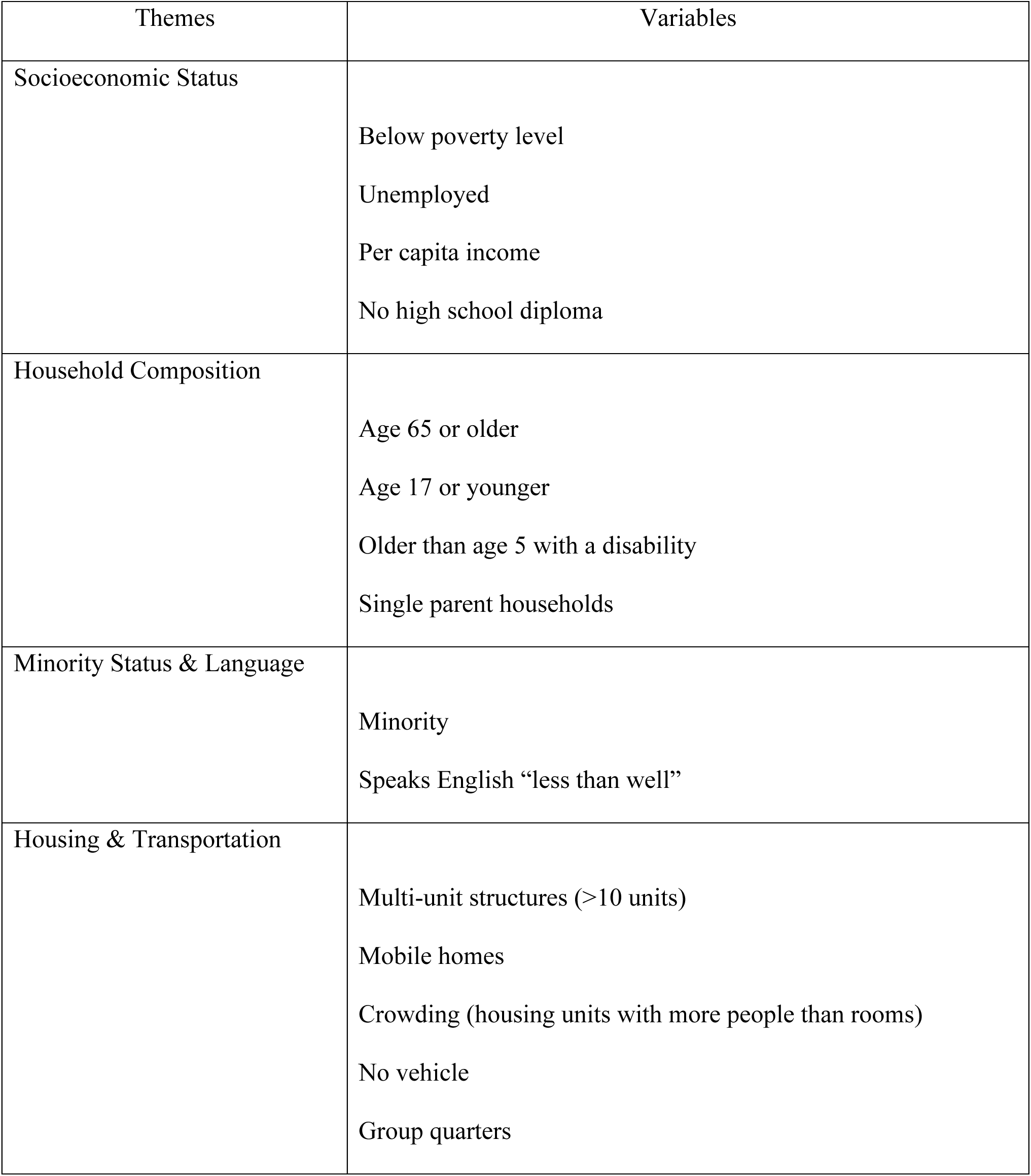
Centers for Disease Control Social Vulnerability Index Themes and Variables

| Themes | Variables |
| --- | --- |
| Socioeconomic Status | Below poverty level<br>Unemployed<br>Per capita income<br>No high school diploma |
| Household Composition | Age 65 or older<br>Age 17 or younger<br>Older than age 5 with a disability<br>Single parent households |
| Minority Status & Language | Minority<br>Speaks English “less than well” |
| Housing & Transportation | Multi-unit structures (>10 units)<br>Mobile homes<br>Crowding (housing units with more people than rooms)<br>No vehicle<br>Group quarters |

To determine subject-level neighborhood SVI, we geocoded each subject’s home address at the time of S1P discharge. Geocoding was conducted using ArcGIS Pro 2.5.1 (Esri, Redlands, CA) and the 2019 Esri Business Analyst composite address locator and street data. Using the geocoded address, we obtained subject census tract as a proxy for neighborhood. The SVI was retrieved from the publicly available website (23). Each subject was assigned the SVI associated with their geocoded census tract at the time of S1P discharge. The SVI is published every few years with each publication representing the census results of the five years prior. SVI data from 2010, 2015, and 2018 SVI data were used for infants born 2007-2012, 2013-2017, and 2018-2020, respectively. Subjects were categorized by SVI tertile: low, middle, and high.

### Data Collection

Medical records were reviewed to abstract detailed information on patient demographics, birth characteristics, S1P operative and discharge characteristics, and S2P admission characteristics (Supplemental Table 1). A health information exchange platform was queried for outside hospital admissions. Echocardiographic variables were obtained by review of reports.

### Outcome Measures

Interstage weight gain, was measured as a continuous variable, average daily weight gain in grams per day, and as a binary variable, interstage growth failure. The primary outcome, average daily weight gain was calculated as an infant’s total change in weight between S1P discharge and S2P admission divided by the duration of their interstage period. The interstage period started on the date of discharge from the S1P admission and concluded on the date of S2P admission. For those that died prior to S2P, the measurement and date of their last documented weight was used to calculate the outcome. The secondary outcome, interstage growth failure, was defined as an average daily weight gain of < 20 grams/day (1,12,24).

### Statistical Analysis

#### Descriptive Statistics

Demographic and clinical characteristics by SVI tertile are summarized using median (interquartile range [IQR]) for continuous variables and frequency (percentage) for categorical variables. Patient characteristics were compared between composite SVI tertiles using the Kruskal-Wallis test for continuous variables and χ2 or Fisher’s exact test for categorical variables. A p-value less than 0.05 was deemed statistically significant.

#### Outcome Analysis

We used multivariable linear regression models to determine whether neighborhood composite SVI was associated with differences in interstage average daily weight gain. To evaluate the relationship of neighborhood SVI and growth failure, we used multivariable logistic regression. The primary exposure, neighborhood SVI, was modeled as a continuous and categorical (tertiles) variable. Model covariates were selected through univariate screening and *a priori* based on clinical knowledge. Final models controlled for: patient race, ethnicity, maternal age, sex, prematurity, presence of a genetic syndrome, birth weight, S1P shunt type [Blalock Thomas Taussig (BTT) versus right ventricle to pulmonary artery conduit (RV-PA)], post-S1P extracorporeal membrane oxygenation, S1P discharge feeding mechanism (oral, oral + nasogastric tube, exclusive gastric or jejunal feeds), tricuspid regurgitation and right ventricular function immediately prior to S1P discharge, interstage digoxin, and year of S1P discharge.

To evaluate whether ISVMP modified the association between composite SVI and weight gain, we introduced an interaction term into our models, SVI*ISVMP, and performed likelihood ratio tests to test the significance of the effect modification. We then calculated the stratum-specific effect of composite SVI by ISVMP group. To determine the relative importance of each of the four SVI themes on our primary outcome, interstage weight gain, we repeated our prior analysis including all four of the SVI themes in the multivariable model (in lieu of the composite SVI variable). To distinguish the effects of ISVMP from secular trends, we performed segmental regression analysis. For this analysis, we fit distinct linear regression models pre- and post-the introduction of the ISVMP and used a t-test to evaluate for a change in intercept by composite SVI tertile.

We performed sensitivity analyses to confirm that the conclusions reached from the main regression models were consistent across different conditions. The first sensitivity analysis excluded patients who were transferred to an outside hospital following S1P but prior to discharge home. The second sensitivity analysis excluded those who died during the interstage period, and the third excluded both those who were transferred and died during the interstage period. There was a small number of infants with missing outcome data (6%). These infants were excluded from our analysis. To ensure that missingness was not differential, SVI and clinical characteristics were compared between infants with and without missing data (Supplemental Table 2).

All statistical analyses used 2-sided tests and were performed using α = 0.05 for the threshold of statistical significance. Stata software (StataCorp, College Station Texas) version 16.1 was used.

## RESULTS

### Patient Characteristics

There was a total of 439 infants, 152 historical controls and 287 ISVMP infants (Figure 1). Overall, 111 infants were excluded because they either died (32 ISVMP and 13 controls), were not discharged from the S1P hospitalization (17 ISVMP and 8 controls), were discharged on palliative care (1 ISVMP and 1 control), underwent biventricular repair (2 ISVMP), were enrolled in an outside home monitoring program (14 ISVMP), resided internationally (3 ISVMP), or had missing data (1 ISVMP and 19 controls, Supplemental Table 2).

**Figure 1.**
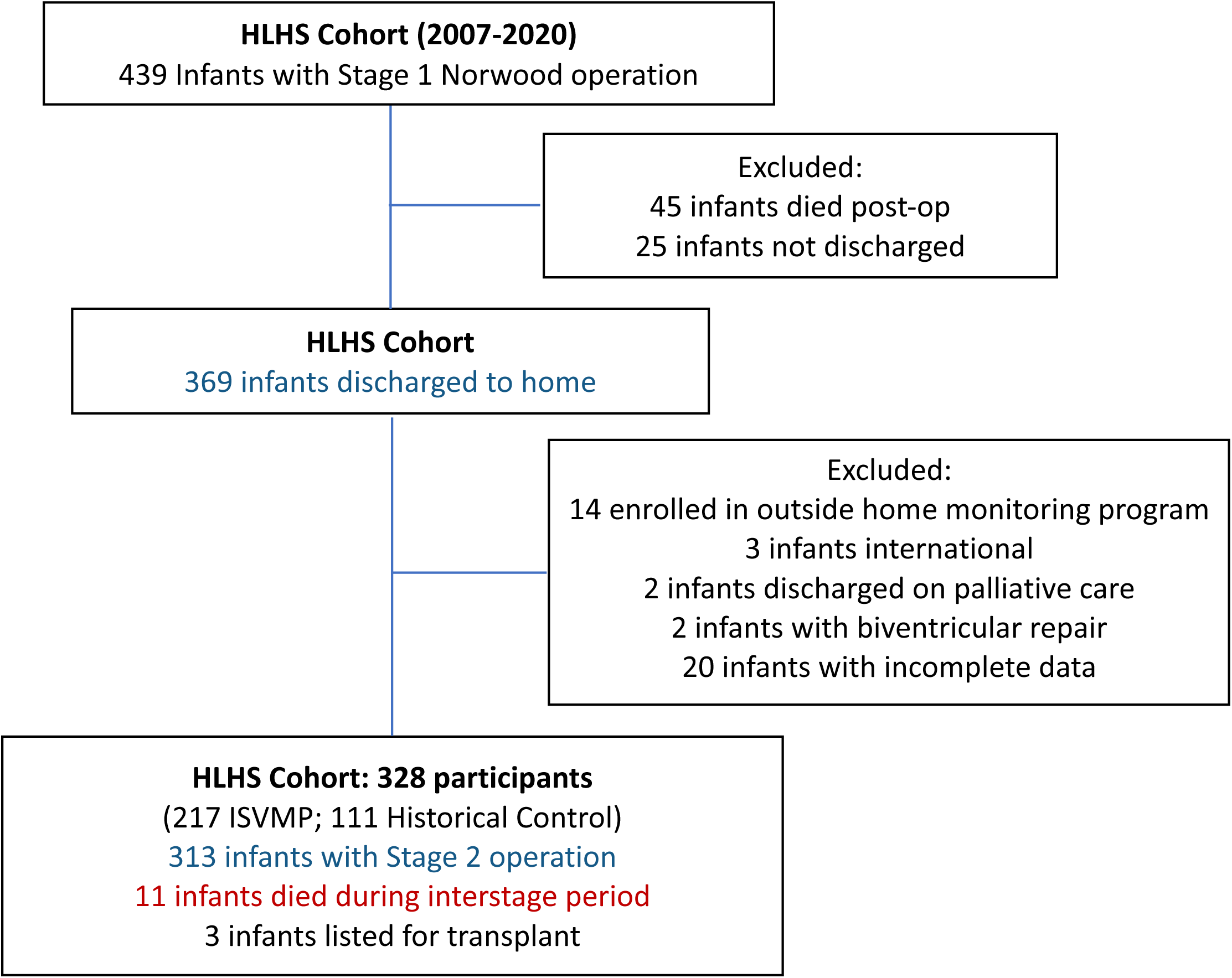
Flow Diagram for Patient Enrollment and Outcomes. Detailed flowchart to describe total infants undergoing surgery, including which participants were included in analysis, and outcomes. ISVMP indicates Infant Single Ventricle Management and Monitoring Program

The composite SVI ranged from 0 to 1 with a median of 0.46. High composite SVI (high vulnerability) infants were born to younger mothers and resided closer to our center than middle and low SVI infants. A greater proportion of infants in the highest SVI tertile were of black race and Hispanic ethnicity (Table 2). There was a clinically insignificant difference in cardiopulmonary bypass times between the tertiles. Otherwise, procedural and post-operative characteries did not differ by SVI tertile (Table 3). At the time of discharge, infants in the highest SVI tertile were more likely to be reliant on supplemental tube feeds. SVI tertiles did not differ in median S1P discharge weight or weight-for-age Z score (Table 4).

**Table 2:** Baseline and Demographic Characteristics WHO WAZ indicates World Health Organization weight-for-age Z score; CHOP, Children’s Hospital of Philadelphia Categorical variables are summarized in count and percentage. Continuous variables are summarized in median and IQR. P values are from χ2 or Fisher’s exact test for categorical variables and Kruskal-Wallis test for continuous variables

|  | Total (n=328) | Low SVI (n=110) | Middle SVI (n=109) | High SVI (n=109) |
| --- | --- | --- | --- | --- |
| Male sex | 204 (62.2) | 74 (67.3) | 62 (56.9) | 68 (62.4) |
| Birth weight, kg | 3.25 (2.90, 3.57) | 3.26 (2.85, 3.59) | 3.27 (3.00, 3.58) | 3.22 (2.80, 3.48) |
| Birth WHO WAZ | -0.14 (-0.95, 0.57) | -0.11 (-1.02, 0.55) | -0.12 (-0.63, 0.62) | -0.22 (-1.17, 0.47) |
| Gestational Age, wks | 39 (38, 39) | 39 (38, 39) | 39 (38, 39) | 39 (38, 39) |
| Maternal Age, yrs* | 29 (24, 34) | 32 (27,35) | 30 (26, 34) | 25 (21, 32) |
| Prenatal diagnosis | 309 (94.2) | 104 (94.5) | 101 (92.7) | 104 (95.4) |
| Race* |  |  |  |  |
| White | 229 (69.8) | 97 (88.2) | 86 (78.9) | 46 (42.2) |
| Black | 48 (14.6) | 5 (4.5) | 7 (6.4) | 36 (33.0) |
| Other | 51 (15.6) | 8 (7.3) | 16 (14.7) | 27 (24.8) |
| Ethnicity* |  |  |  |  |
| Non-Hispanic | 282 (86.0) | 104 (94.5) | 95 (87.2) | 83 (76.1) |
| Hispanic | 42 (12.8) | 4 (3.6) | 12 (11.0) | 26 (23.9) |
| Unknown | 4 (1.2) | 2 (1.8) | 2 (1.8) | 0 (0.0) |
| Distance from<br>CHOP, miles* | 52.8 (20.2, 90.6) | 57.4 (31.9, 97.7) | 57.3 (20.1, 105.9) | 46.9 (12.5, 80.7) |
| Genetic syndrome | 16 (4.9) | 9 (8.2) | 2 (1.8) | 5 (4.6) |

\*Statistically significant difference between SVI tertiles

**Table 3:** Stage 1 Operative Characteristics BTT indicates Blalock-Thomas-Taussig shunt; RV-PA, right ventricle to pulmonary artery; CPR, cardiopulmonary resuscitation; ECMO, extracorporeal membrane oxygenation Categorical variables are summarized in count and percentage. Continuous variables are summarized in median and IQR. P values are from χ2 or Fisher’s exact test for categorical variables and Kruskal-Wallis test for continuous variables

|  | Total<br>(n=328) | Low SVI<br>(n=110) | Middle SVI<br>(n=109) | High SVI<br>(n=109) |
| --- | --- | --- | --- | --- |
| Age at Stage 1, days | 4 (2, 6) | 4 (2, 6) | 4 (3, 6) | 4 (2, 6) |
| Stage 1 palliation |  |  |  |  |
| BTT shunt | 182 (55.5) | 52 (47.3) | 67 (61.5) | 63 (57.8) |
| RV-PA conduit | 141 (43.0) | 56 (50.9) | 42 (38.5) | 43 (39.4) |
| BTT + RV-PA conduit | 5 (1.5) | 2 (1.8) | 0 (0.0) | 3 (2.9) |
| CPB, min* | 86 (77, 102) | 84 (76, 95) | 84 (78, 103) | 91 (80, 110) |
| Mechanical Ventilation, days | 2.5 (1, 5) | 3 (1, 5) | 2 (1, 5) | 3 (1,5) |
| CPR | 31 (9.5) | 11 (10.0) | 7 (6.4) | 13 (11.9) |
| ECMO | 18 (5.5) | 4 (3.6) | 6 (5.5) | 8 (7.3) |
| Length of hospital stay, days | 26 (19, 40) | 23 (18, 33) | 27 (19, 43) | 28 (20, 41) |

\*Statistically significant difference between SVI tertiles

**Table 4:** Stage 1 Discharge Characteristics WHO WAZ indicates World Health Organization weight-for-age Z score; SVI, social vulnerability index; PO, per os; NGT, nasogastric tube Categorical variables are summarized in count and percentage. Continuous variables are summarized in median and IQR. P values are from χ2 or Fisher’s exact test for categorical variables and Kruskal-Wallis test for continuous variables

|  | Total<br>(n=328) | Low SVI<br>(n=110) | Middle SVI<br>(n=109) | High SVI<br>(n=109) |
| --- | --- | --- | --- | --- |
| Weight, kg | 3.49<br>(3.16, 3.87) | 3.49<br>(3.11, 3.85) | 3.57<br>(3.24, 3.91) | 3.45<br>(3.15, 3.90) |
| WHO WAZ | -1.36<br>(-2.03, -0.72) | -1.47<br>(-2.2, -0.77) | -1.35<br>(-1.86, -0.69) | -1.24<br>(-2.15, -0.75) |
| SVI Composite* | 0.46 (0.24, 0.70) | 0.17 (0.08, 0.24) | 0.46 (0.37, 0.54) | 0.80 (0.71, 0.92) |
| Socioeconomic Status | 0.46 (0.25, 0.70) | 0.19 (0.09, 0.33) | 0.47 (0.33, 0.60) | 0.79 (0.68, 0.94) |
| Household Composition | 0.47 (0.26, 0.72) | 0.27 (0.16, 0.44) | 0.42 (0.26, 0.58) | 0.79 (0.62, 0.91) |
| Minority Status & Language | 0.49 (0.26, 0.74) | 0.29 (0.15, 0.45) | 0.41 (0.26, 0.66) | 0.78 (0.63, 0.89) |
| Housing & Transportation | 0.44 (0.23, 0.68) | 0.18 (0.08, 0.26) | 0.51 (0.35, 0.63) | 0.73 (0.55, 0.86) |
| Prescribed digoxin | 147 (47.9) | 56 (50.9) | 51 (46.8) | 50 (45.9) |
| Feeding mechanism* |  |  |  |  |
| PO | 83 (25.3) | 34 (30.9) | 36 (33.0) | 13 (11.9) |
| PO + NGT | 193 (58.8) | 58 (52.7) | 62 (56.9) | 73 (67.0) |
| Exclusive tube feeds | 52 (15.9) | 18 (16.4) | 11 (10.1) | 23 (21.1) |
| Ventricular function |  |  |  |  |
| Normal to mildly diminished | 315 (96.0) | 108 (98.2) | 101 (92.7) | 106 (97.2) |
| Moderate to severely diminished | 13 (4.0) | 2 (1.8) | 8 (7.3) | 3 (2.8) |
| AV valve regurgitation |  |  |  |  |
| None-Mild | 270 (82.3) | 94 (85.5) | 92 (84.4) | 84 (77.1) |
| Moderate-severe | 58 (17.7) | 16 (14.5) | 17 (15.6) | 25 (22.9) |

\*Statistically significant difference between SVI tertiles

As composite SVI tertile increased, interstage average daily weight gain decreased, and the frequency of growth failure increased (Table 5). Adjusted analysis demonstrated that composite SVI was significantly associated with interstage weight gain and growth failure. High SVI infants gained 3 gm/day less and were over three times as likely to experience interstage growth failure compared to low SVI infants (adjusted OR 3.26; 95% CI 1.59-6.72; P = 0.001). Infants enrolled in ISVMP gained nearly 2 gm/day more than historical controls and were half as likely to develop growth failure (adjusted OR 0.48, 95% CI 0.27, 0.85; P = 0.01).

**Table 5:** Interstage Outcomes ISVMP indicates infant single-ventricle management and monitoring program; WHO WAZ, World Health Organization weight-for-age Z score Categorical variables are summarized in count and percentage. Continuous variables are summarized in median and IQR. P values are from χ2 or Fisher’s exact test for categorical variables and Kruskal-Wallis test for continuous variables

|  | Total<br>(n=328) | Low SVI<br>(n=110) | Middle SVI<br>(n=109) | High SVI<br>(n=109) | P-Value |
| --- | --- | --- | --- | --- | --- |
| Enrolled in ISVMP | 217 (66.2) | 78 (70.9) | 72 (66.1) | 67 (61.5) | 0.35 |
| Interstage duration, days | 108 (86, 129) | 102 (84, 124) | 113 (91, 127) | 112 (86, 139) | 0.13 |
| Pre-Stage 2 Weight, kg | 5.96<br>(5.36, 6.60) | 5.90<br>(5.36, 6.40) | 6.10<br>(5.64, 6.70) | 5.81<br>(5.30, 6.60) | 0.09 |
| Pre-Stage 2 WHO WAZ | -1.51<br>(-2.13, -0.77) | -1.46<br>(-2.13, -0.77) | -1.23<br>(-1.77, -0.52) | -1.56<br>(-2.25, -0.86) | 0.05 |
| Daily weight gain, gm/day | 22 (18, 26) | 23 (20, 28) | 21 (18, 25) | 20 (17, 25) | 0.004 |
| Growth Failure<sup>1</sup> | 133 (40.7) | 31 (28.4) | 44 (40.4) | 58 (53.2) | 0.001 |
| Interstage mortality | 11 (3.4) | 4 (3.6) | 5 (4.6) | 2 (1.8) | 0.56 |

<sup>1</sup>Growth failure was defined as an average daily weight gain of < 20 grams/day

### Effect Modification by ISVMP

The interaction between neighborhood composite SVI and ISVMP was significant such that enrollment in ISVMP strongly attenuated the effect of neighborhood SVI tertile on interstage weight gain and growth failure (Table 6; Figure 2).High and middle SVI infants enrolled in ISVMP were one quarter as likely to develop interstage growth failure compared to those not enrolled in ISVMP (High SVI x ISVMP interaction: adjusted OR 0.26; 95% CI 0.07-0.98; P = 0.042; Middle SVI x ISVMP interaction: adjusted OR 0.24; 95% CI 0.06-0.93; P = 0.04; Supplemental Table 3). Low SVI infants did not demonstrate the same benefit from enrollment in ISVMP (Table 6; Supplemental Table 3).

**Figure 2.**
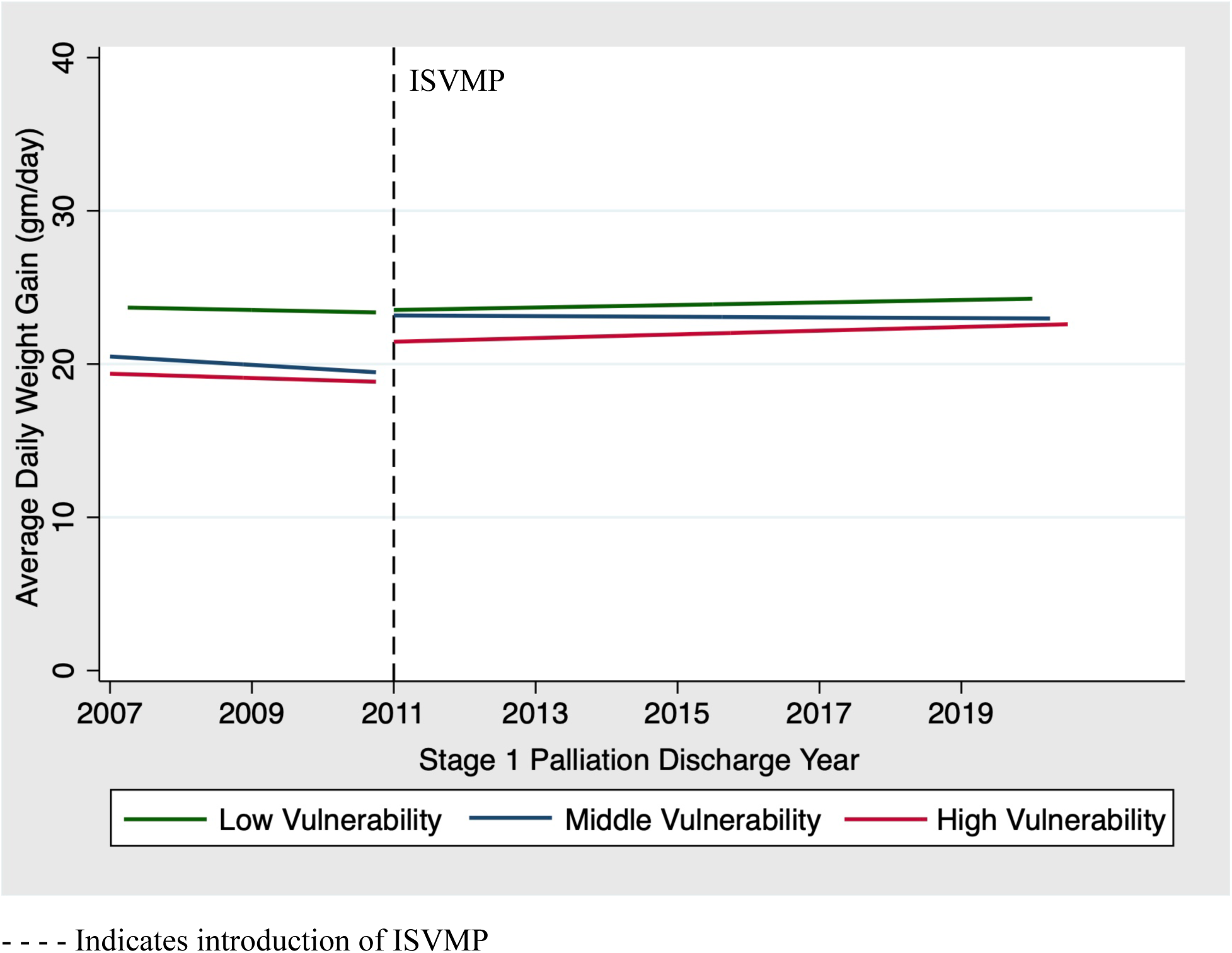
Segmental Regression Analysis of Daily Weight Gain by SVI Tertile over Time. The introduction of the ISVMP was associated with a significant change in the daily weight gain of the middle SVI tertile (+ 3.8 grams/day; 95% CI 0.69-6.86; P = 0.02). There was an increase in the daily weight gain of the high SVI tertile, however, this change did not reach statistical significance (+ 2.2 grams/day; 95% CI -0.36-4.77; P = 0.09). The low SVI tertile did not demonstrate any change in daily weight gain with the introduction of the ISVMP.

**Table 6:** Multivariable Analysis with Interaction Term for Outcome Daily Weight Gain ISVMP indicates infant single-ventricle management and monitoring program; S1P, stage 1 palliation; BTT; Blalock–Thomas–Taussig shunt; RV-PA, right ventricle to pulmonary artery; CPR, cardiopulmonary resuscitation; ECMO, extracorporeal membrane oxygenation; PO, per os; NGT, nasogastric tube

|  | Daily Weight Gain (gm/day) |  |
| --- | --- | --- |
|  | Weight Differential<br>(95% CI) | P-value |
| Middle vs Low Tertile | -4.57 (-7.63, -1.49) | 0.004 |
| High vs Low Tertile | -5.28 (-8.41, -2.16) | 0.001 |
| ISVMP | -0.79 (-3.56, 1.98) | 0.57 |
| Middle x ISVMP interaction | 4.00 (0.30, 7.71) | 0.03 |
| High x ISVMP interaction | 3.44 (-0.19, 7.09) | 0.06 |
| Race |  |  |
| White | Reference |  |
| Black | 0.66 (-1.62, 2.96) | 0.57 |
| Other | 0.48 (-1.85, 2.81) | 0.40 |
| Hispanic ethnicity | 0.06 (-2.51, 2.64) | 0.96 |
| Maternal age | -0.05 (-0.17, 0.07) | 0.39 |
| Prematurity | -0.20 (-3.16, 2.75) | 0.89 |
| Birth weight | -0.08 (-1.63, 1.46) | 0.91 |
| Female gender | -2.30 (-3.78, -0.81) | 0.003 |
| Genetic syndrome | -1.42 (-4.81, 1.98) | 0.41 |
| S1P |  |  |
| BTT shunt | Reference |  |
| RV-PA conduit | 1.20 (-0.36, 2.76) | 0.13 |
| BTT + RV-PA conduit | 0.60 (-5.27, 6.47) | 0.84 |
| S1P ECMO | 1.17 (-2.02, 4.37) | 0.47 |
| S1P prescribed digoxin | -0.97 (-2.47, 0.54) | 0.21 |
| S1P Discharge Feeding Mechanism |  |  |
| Exclusive PO feeds | Reference |  |
| PO + NGT | -1.08 (-2.79, 0.63) | 0.22 |
| Exclusive enteral feeds | -3.11 (-5.49, -0.74) | 0.01 |
| S1P Discharge Ventricular Function |  |  |
| Normal to mildly diminished | Reference |  |
| Moderate to severely diminished | -1.22 (-4.84, 2.40) | 0.50 |
| AV valve regurgitation |  |  |
| None - mild | Reference |  |
| Moderate - severe | -0.74 (-2.61, 1.12) | 0.43 |

Stratified analysis demonstrated that historical control infants in the highest and middle SVI tertiles gained 4 gm/day less and were greater than seven times as likely to experience interstage growth failure than those in the lowest SVI tertile (high vs. low: adjusted OR 12.5; 95% CI 2.5-62.2; P = 0.002; middle vs. low: adjusted OR 7.8; 95% CI 2.0-31.2; P = 0.004; Table 7). For every 0.1 increase in the neighborhood composite SVI, the risk of growth failure increased by 45% (adjusted OR 1.45; 95% CI 1.15-1.81; P = 0.001). In the historical control cohort, discharge on a combination of oral and nasogastric feeds compared to exclusive oral feeds was identified as additional risk for growth failure (adjusted OR 5.42; 95% CI 1.47-20.0; P = 0.01). Sensitivity analysis yielded comparable results (Supplemental Table 4).

**Table 7:**
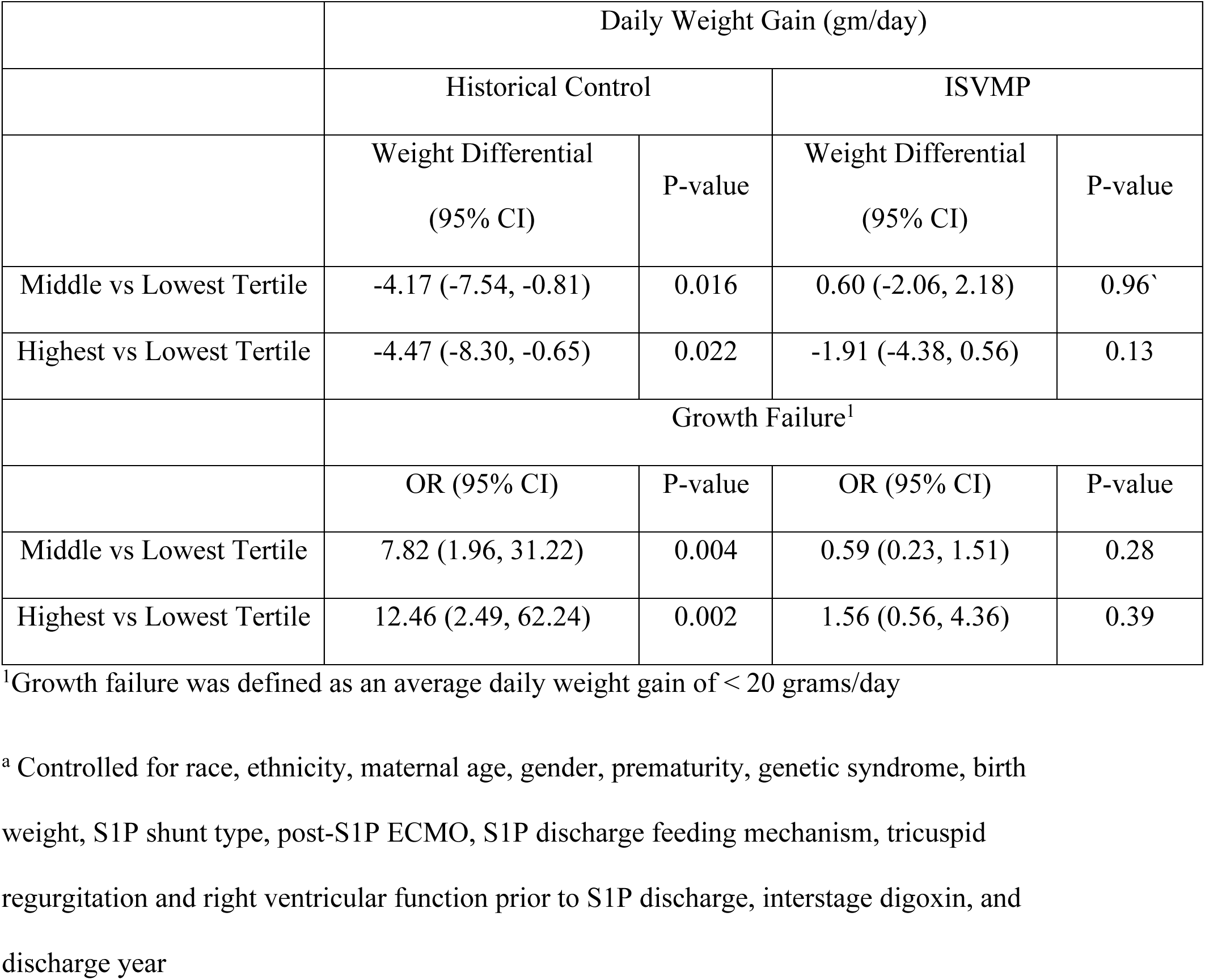
Multivariable Analysis Stratified by ISVMP ISVMP indicates infant single-ventricle management and monitoring program

|  | Daily Weight Gain (gm/day) |  |  |  |
| --- | --- | --- | --- | --- |
|  | Historical Control |  | ISVMP |  |
|  | Weight Differential<br>(95% CI) | P-value | Weight Differential<br>(95% CI) | P-value |
| Middle vs Lowest Tertile | -4.17 (-7.54, -0.81) | 0.016 | 0.60 (-2.06, 2.18) | 0.96<sup>c</sup> |
| Highest vs Lowest Tertile | -4.47 (-8.30, -0.65) | 0.022 | -1.91 (-4.38, 0.56) | 0.13 |
|  | Growth Failure<sup>1</sup> |  |  |  |
|  | OR (95% CI) | P-value | OR (95% CI) | P-value |
| Middle vs Lowest Tertile | 7.82 (1.96, 31.22) | 0.004 | 0.59 (0.23, 1.51) | 0.28 |
| Highest vs Lowest Tertile | 12.46 (2.49, 62.24) | 0.002 | 1.56 (0.56, 4.36) | 0.39 |

<sup>1</sup>Growth failure was defined as an average daily weight gain of < 20 grams/day
<sup>a</sup> Controlled for race, ethnicity, maternal age, gender, prematurity, genetic syndrome, birth weight, S1P shunt type, post-S1P ECMO, S1P discharge feeding mechanism, tricuspid regurgitation and right ventricular function prior to S1P discharge, interstage digoxin, and discharge year

After the introduction of the ISVMP, there was no longer a significant difference in interstage weight gain or growth failure by SVI tertile (Table 7). However, high SVI infants gained 2 gm/day less than their middle and high SVI counterparts. In infants enrolled in ISVMP, significant risk factors for growth failure were female sex (adjusted OR 2.5; 95% CI 1.13-5.53; P = 0.02), discharge on exclusive tube feeds (adjusted OR 6.8; 95% CI 1.89-24.2; P = 0.003), and BTT shunt as compared to RV-PA conduit (adjusted OR 3.0; 95% CI 1.30-7.15; P = 0.01). Sensitivity analyses did not significantly alter these results (Supplemental Table 4).

SVI themes were only modestly correlated (Pearson’s R = 0.3-0.7). Multivariable models including the four SVI themes, demonstrated that in the historical control cohort “housing type and transportation” was the only SVI theme associated with interstage weight gain. For every 0.1 increase in the “housing type and transportation” SVI, an infant gained 0.6 gm/day less during the interstage period (-0.59 gm/day; 95% CI -1.14, -0.4; P = 0.035). Infants in the highest and middle SVI “housing type and transportation” tertiles gained 4 gm/day less than infants in the lowest SVI tertile (high vs. low: -4.11 gm/day; 95% CI -7.95, -0.27; P = 0.036; middle vs. low; - 4.53 gm/day; 95% CI -8.15, -0.92; P = 0.014). After the introduction of the ISVMP, there was no significant difference in interstage weight gain by any of the SVI themes.

### Segmental Regression Analysis

On adjusted segmental regression analysis, we found that the introduction of ISVMP was associated with a significant change in the daily weight gain of the middle SVI tertile (change of + 3.8 grams/day; 95% CI 0.69-6.86; P = 0.02, Figure 2) suggesting that our findings are unlikely to be the consequence of secular trends. There was an increase in the daily weight gain of the high SVI tertile, however, this change did not reach statistical significance (change of + 2.2 grams/day; 95% CI -0.36-4.77; P = 0.09, Figure 2). The low SVI tertile did not demonstrate any change in daily weight gain with the introduction of the ISVMP.

## DISCUSSION

In this study, we investigated the impact of a home monitoring program on the association of neighborhood social vulnerability and weight gain among infants with HLHS in the interstage period. We found that high neighborhood social vulnerability was strongly associated with poor weight gain and growth failure. Enrollment in ISVMP largely attenuated this association such that for infants enrolled in ISVMP, social vulnerability was no longer a risk factor for poor weight gain. This is the first study to report a significant impact of home monitoring on disparities in interstage weight gain and growth failure.

We found that in the era prior to home monitoring, small increases in neighborhood social vulnerability particularly in “housing type and transportation” significantly increased an infant’s risk of growth failure after S1P discharge. Consistent with our findings, prior studies have reported an association between social factors and poor outcomes following S1P. Secondary analysis of data from the Single Ventricle Reconstruction (SVR) Trial identified a significant association between low neighborhood SES (high vulnerability) and worse 1-year transplant-free survival following S1P (2). Low SES has also been associated with longer hospital length of stay during S1P, as well as total hospitalized days over the first year of life (25). In addition to SES, Hispanic race, birth to a teenage mother and residing in a home with a single caregiver have been identified as demographic and social risk factors for interstage mortality (3,4).

To our knowledge, our study is the first to investigate the impact of neighborhood social vulnerability on infants with HLHS. The SVI is a more comprehensive measure of social determinants of health then traditional SES scales (18). It highlights many of the social factors that can limit access to and utilization of healthcare. Thus, the independent association of neighborhood social vulnerability and interstage weight gain is not unexpected. Infants with HLHS require specialized and frequent follow-up to ensure adequate weight gain after discharge (1). Based on our findings, we speculate that infants with a high SVI face greater barriers to follow-up care due to inaccessibility of transportation, competing financial needs, job-related stress, and limited caregiver support. Infants with HLHS often require fortified formula and supplemental tube feeds to achieve adequate weight gain (26,27). In high vulnerability infants, inadequate education, low health literacy and English language insufficiency may present obstacles to accurate formula mixing (28–30). Additionally, adherence to supplemental tube feeds may be influenced by caregiver support and access to transportation if a caregiver requires assistance replacing the dislodged tube.

We found that the introduction of ISVMP markedly reduced disparities in interstage growth failure. In middle and high vulnerability infants, enrollment in ISVMP was associated with a dramatic improvement in interstage weight gain. Caregiver education, close communication, and easy access to the healthcare system are essential components of home monitoring and may be particularly beneficial to infants with high vulnerability. In preparation for discharge, parents receive intensive and personalized education on equipment use (pulse oximeter, weight scale, nasogastric tube), daily measurements, data interpretation, and red flags (1). Prior to S1P discharge, the ISVMP requires parents to room-in for 24 hours and independently perform all aspects of care. Upon discharge, parents are encouraged to communicate frequently with the home monitoring team. Families have easy access to a dedicated ISVMP nurse practitioner during regular working hours and the on-call cardiology team is readily available after hours. Social work resources and more recently telemedicine can aid families with difficulties accessing transportation (31). If concerns arise over home surveillance parameters or lapses in follow-up care, families may be instructed to present for hospital readmission.

Female sex, discharge from the S1P on supplemental tube feeds, and presence of a BTT shunt were identified as independent predictors of interstage growth failure in infants enrolled in ISVMP. These findings are largely consistent with those of prior studies (10,12,32). Tube feeds (in contrast to exclusive oral feeds) are likely a surrogate measure for increased complexity of the neonatal hospitalization. Secondary analysis of data from the SVR trial found that infants with a BTT shunt had poorer weight gain than those with an RV-PA conduit (10). Burch *et al.* hypothesizes that this may be explained by the higher energy expenditure associated with BTT shunt physiology (10,33,34). Furthermore, a recent pilot study in shunt-dependent infants demonstrated an inverse relationship between brain natriuretic peptide (BNP) and insulin like growth factor 1, a correlate of weight gain in infancy (35,36). Given variability in right ventricle volume loading by shunt type and size, the relationship of shunt type, BNP, and weight gain merits further investigation.

Our study has important implications for efforts to eliminate disparities in pediatric cardiac outcomes. While disparities in this population have significantly improved with the adoption of home monitoring, high SVI infants continue to lag behind their middle and low SVI peers in interstage weight gain. A recent study by Jackson *et al*. identified lack of English language fluency, lower median household income, and Medicaid insurance as risk factors for lower caregiver adherence to mobile health interstage home monitoring (37). Qualitative studies could provide valuable insight into these obstacles to adherence and inspire alternative strategies to address disparities. In addition, the discontinuation of home monitoring following S2P marks a major transition point for infants with HLHS. Some caregivers express feelings of stress and anxiety around caring for their infants without the aid of home monitoring (38). It is unknown whether these feelings may be heightened in high vulnerability families. Thus, future efforts investigating social vulnerability following S2P are warranted.

### Limitations

We acknowledge several limitations of our study. Our study design is retrospective with a “before versus after” analysis. As such, we adjusted for a broad range of covariates including those with temporal variability (discharge on digoxin, shunt type, discharge year). To distinguish the effects of ISVMP from secular trends, we used segmental regression analysis. Despite this, we recognize that subtle practice changes could introduce unmeasured confounding. Secondly, this is a single center study at a large academic medical center. While the study population is diverse and likely representative of other high volume academic centers, our results may be less generalizable to smaller volume programs or those with different models of home monitoring.

## CONCLUSION

In this single center study, we found that in infants with HLHS, increased social vulnerability is a risk factor for poor interstage weight gain. Enrollment in ISVMP significantly reduced growth disparities, but high vulnerability infants continued to gain less weight than their middle and low vulnerability peers. These findings highlight the need for additional research to identify novel strategies to address disparities. Given the marked effect of home monitoring, further investigation into disparities following discontinuation of these programs following S2P are warranted.

## Data Availability

The data, analytic methods, and study materials will not be made available to other researchers for purposes of reproducing the results or replicating the procedure.

## Abbreviations and Acronyms

BTT: Blalock-Thomas-Taussig
HLHS: Hypoplastic Left Heart Syndrome
IQR: Interquartile Range
ISVMP: Infant Single Ventricle Management & Monitoring Program
OR: Odds Ratio
RV-PA: Right ventricle to pulmonary artery
S1P: Stage 1 Norwood palliation
S2P: Stage 2 palliation
SVR: Single Ventricle Reconstruction

## ACKNOWLEDGEMENTS

This work was supported by the Cardiac Center Clinical Research Core at the Children’s Hospital of Philadelphia

## CLINICAL PERSPECTIVE

<u>What is New?</u>

- This study is the first to report a significant impact of home monitoring on disparities in interstage weight gain and growth failure.
- High neighborhood social vulnerability is a risk factor for poor interstage weight gain.
- Home monitoring can significantly decrease interstage growth disparities, however, high vulnerability infants continue to lag behind their middle and low vulnerability peers.

<u>What are the Clinical Implications?</u>

- Interstage care should focus on assessing and addressing socioeconomic risk factors particularly those that may affect access to care.
- Additional studies are needed to identify obstacles to care in high vulnerability infants and evaluate disparities following discontinuation of home monitoring following Stage 2 palliation.

## SUPPLEMENTAL TABLES

**Table 1:** Study Covariates S1P indicates Stage 1 Norwood palliation; HLHS, hypoplastic left heart syndrome; BTT, Blalock-Taussig-Thomas; RV-PA, right ventricle-pulmonary artery; CPR, cardiopulmonary resuscitation; ECMO; extracorporeal membrane oxygenation; S2P, Stage 2 palliation.

| Demographics |  |
| --- | --- |
|  | Sex<br>Race<br>Ethnicity<br>Gestational Age<br>Maternal Age<br>Age at S1P<br>Birth weight-for-age Z score<br>Presence of genetic syndrome<br>Presence of extra-cardiac anomaly<br>Pre vs post-natal diagnosis |
| Anatomic and S1P Characteristics |  |
|  | HLHS subtype<br>Presence of heterotaxy<br>Presence of pulmonary vein anomalies<br>Need for catheter or surgical intervention pre-S1P<br>Shunt type – BTT or RV-PA shunt<br>Cardiopulmonary bypass time<br>Duration of mechanical ventilation (days)<br>Need for post-operative CPR or ECMO<br>Presence of post-operative arrhythmia |
| S1P Discharge Characteristics |  |
|  | Length of stay (days)<br>Weight-for-age Z score<br>Number of discharge medications<br>Prescription for digoxin<br>Mechanism of feeding (oral, oral + tube, tube)<br>Presence of tricuspid regurgitation and ventricular dysfunction on pre-discharge echocardiogram<br>Distance of primary residence from CHOP (miles) |
| S2P Characteristics |  |
|  | Length of stay (days)<br>Weight-for-age Z score<br>Number of discharge medications<br>Prescription for digoxin<br>Mechanism of feeding (oral, oral + tube, tube)<br>Presence of tricuspid regurgitation and ventricular dysfunction on pre-discharge echocardiogram<br>Distance of primary residence from CHOP (miles)<br>Map coordinates of primary residence |

**Table 2:**
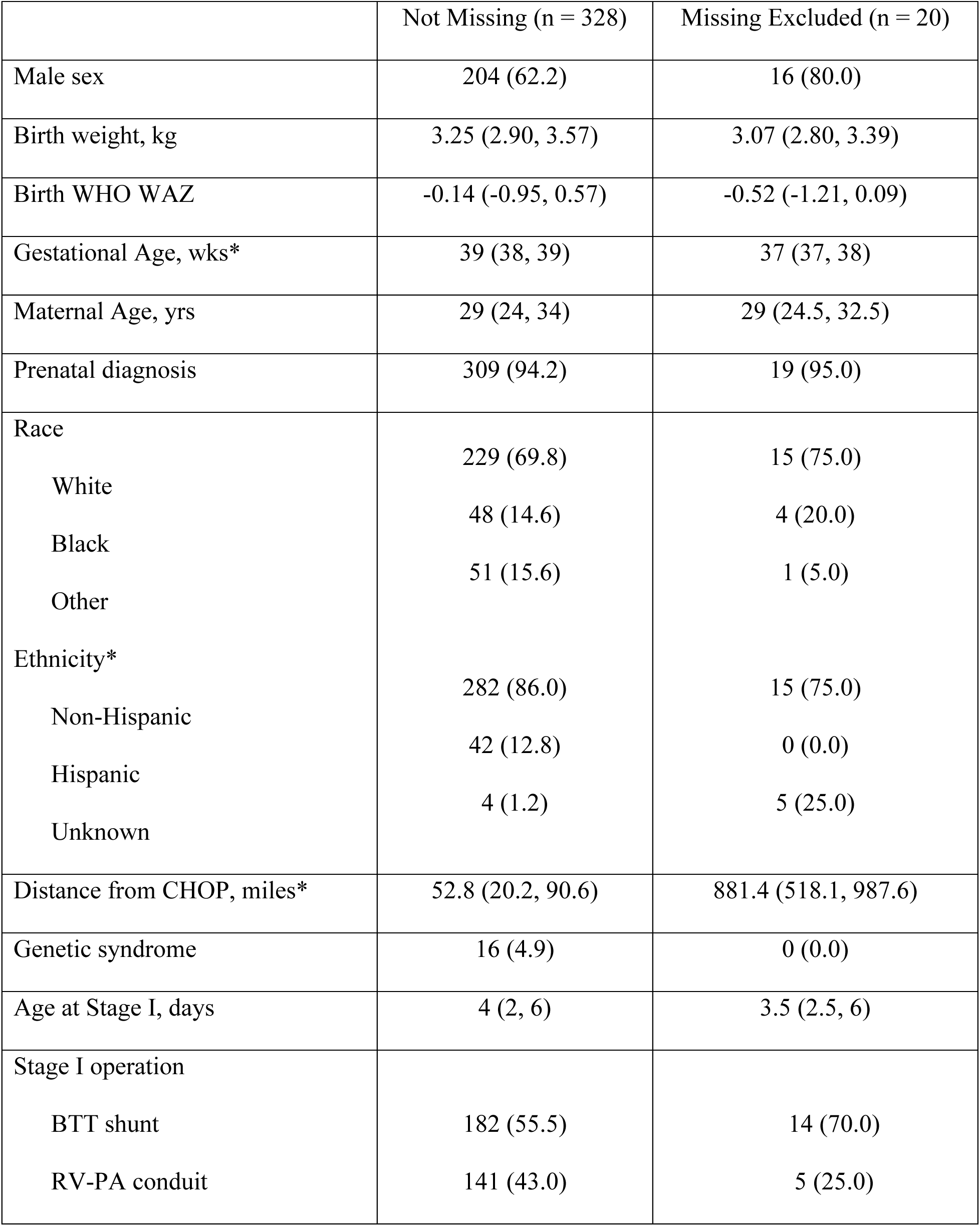
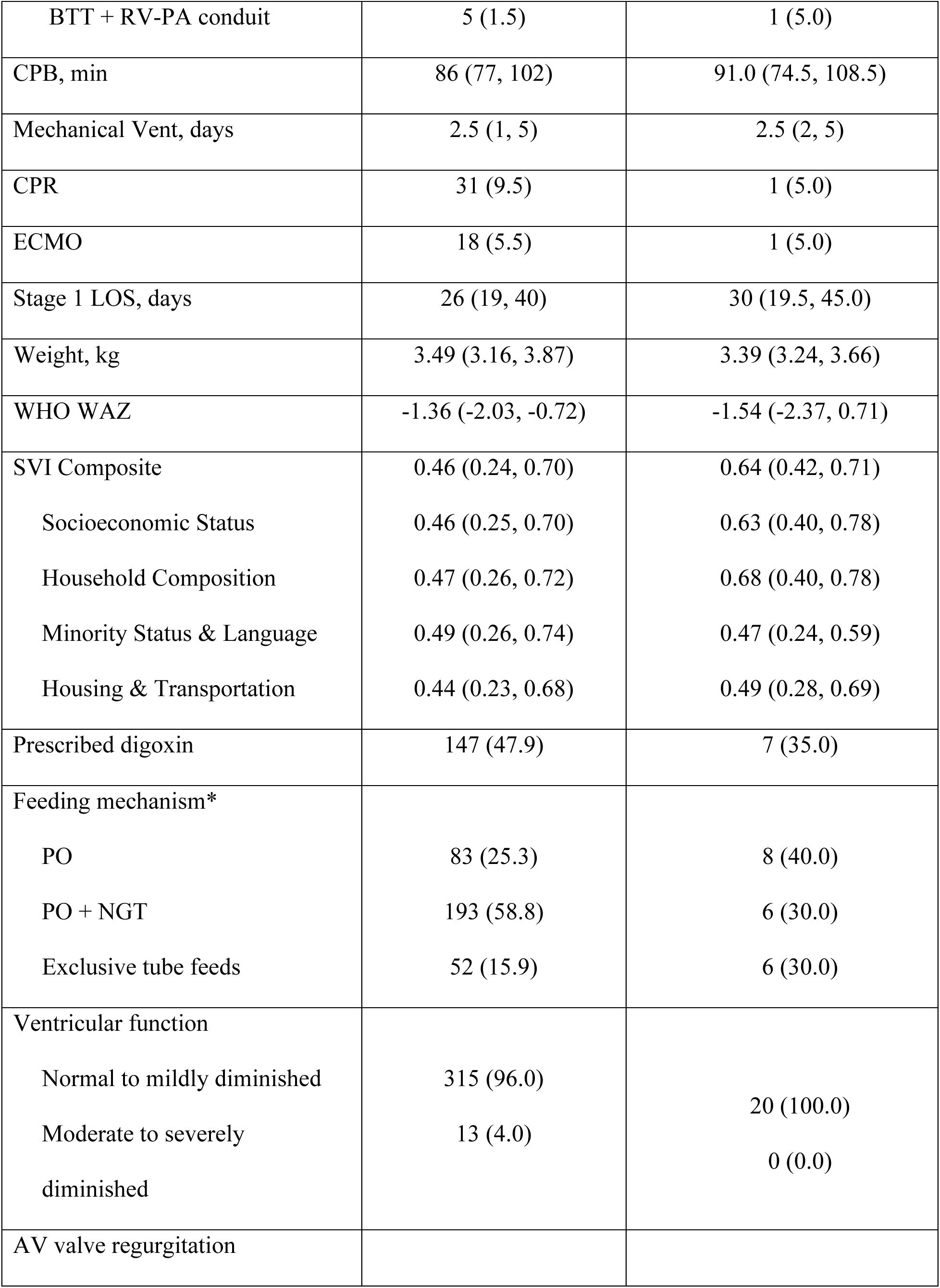
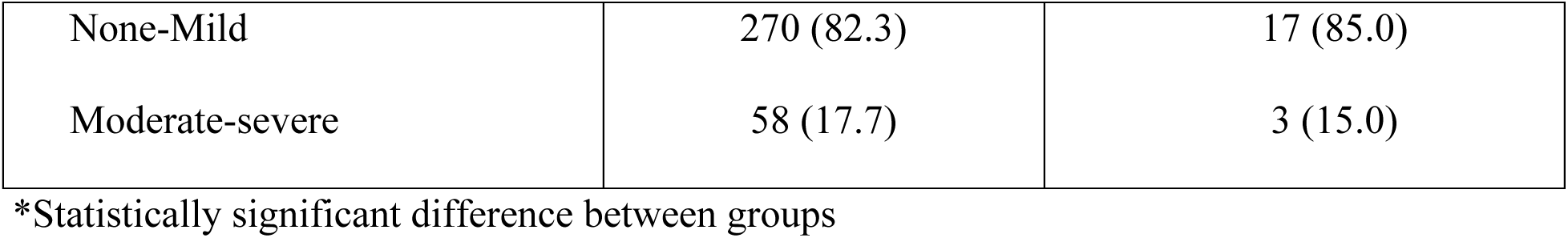
Demographic, Operative, and Discharge Characteristics by Missingness

**Table 3:** Multivariable Analysis with Interaction Term for Outcome Growth Failure ISVMP indicates infant single-ventricle management and monitoring program; S1P, stage 1 palliation; BTT; Blalock-Thomas-Taussig shunt; RV-PA, right ventricle to pulmonary artery; CPR, cardiopulmonary resuscitation; ECMO, extracorporeal membrane oxygenation; PO, per os; NGT, nasogastric tube

|  | Growth Failure<sup>1</sup> |  |
| --- | --- | --- |
|  | OR (95% CI) | P-value |
| Middle vs Low Tertile | 4.58 (1.51, 13.97) | 0.007 |
| High vs Low Tertile | 7.91 (2.49, 25.1) | 0.000 |
| ISVMP | 1.30 (0.46, 3.68) | 0.62 |
| Middle x ISVMP interaction | 0.24 (0.06, 0.93) | 0.039 |
| High x ISVMP interaction | 0.26 (0.07, 0.98) | 0.042 |
| Race |  |  |
| White | Reference |  |
| Black | 0.81 (0.36, 1.82) | 0.61 |
| Other | 0.41 (0.16, 1.00) | 0.05 |
| Hispanic ethnicity | 1.82 (0.71, 4.64) | 0.21 |
| Maternal age | 1.02 (0.97, 1.06) | 0.45 |
| Prematurity | 0.59 (0.20, 1.78) | 0.35 |
| Birth weight | 1.23 (0.70, 2.15) | 0.47 |
| Female gender | 1.94 (1.14, 3.29) | 0.01 |
| Genetic syndrome | 2.53 (0.70, 9.15) | 0.16 |
| S1P |  |  |
| BTT shunt | Reference |  |
| RV-PA conduit | 0.54 (0.31, 0.94) | 0.03 |
| BTT + RV-PA conduit | 0.57 (0.08, 4.20) | 0.59 |
| S1P ECMO | 1.65 (0.55, 4.96) | 0.38 |
| S1P prescribed digoxin | 1.26 (0.73, 2.17) | 0.40 |
| S1P discharge feeding mechanism |  |  |
| Exclusive PO feeds | Reference |  |
| PO + NGT | 2.09 (1.10, 3.99) | 0.03 |
| Exclusive tube feeds | 2.63 (1.12, 6.20) | 0.03 |
| S1P Discharge Ventricular Function |  |  |
| Normal to mildly diminished | Reference |  |
| Moderate to severely diminished | 1.46 (0.43, 5.00) | 0.54 |
| AV valve regurgitation |  |  |
| None - mild | Reference |  |
| Moderate - severe | 1.00 (0.52, 1.96) | 0.99 |

<sup>1</sup>Growth failure was defined as an average daily weight gain of < 20 grams/day

**Table 4:** Multivariable Analysis Stratified by ISVMP: Results of Sensitivity Analyses ISVMP indicates infant single-ventricle management and monitoring program; S1P, Stage 1 palliation; ECMO, extracorporeal membrane oxygenation

| **Sensitivity Analysis #1: Non-Transfer (n= 313)** |  |  |  |  |
| --- | --- | --- | --- | --- |
|  | Daily Weight Gain (gm/day) |  |  |  |
|  | Historical Control |  | ISVMP |  |
|  | Weight Differential (95% CI) | P-value | Weight Differential (95% CI) | P-value |
| Middle vs Low Tertile | -3.65 (-7.08, -0.22) | 0.03 | 0.23 (-1.89, 2.34) | 0.83 |
| High vs Low Tertile | -4.88 (-8.97, -0.79) | 0.02 | -1.73 (-4.20, 0.74) | 0.17 |
|  | Growth Failure |  |  |  |
|  | OR (95% CI) | P-value | OR (95% CI) | P-value |
| Middle vs Low Tertile | 5.97 (1.45, 24.51) | 0.01 | 0.59 (0.23, 1.51) | 0.27 |
| High vs Low Tertile | 12.65 (2.14, 74.7) | 0.005 | 1.56 (0.56, 4.35) | 0.40 |
| **Sensitivity Analysis #2: Survived to S2P (n = 312)** |  |  |  |  |
|  | Daily Weight Gain (gm/day) |  |  |  |
|  | Historical Control |  | ISVMP |  |
|  | Weight Differential (95% CI) | P-value | Weight Differential (95% CI) | P-value |
| Middle vs Low Tertile | -4.54 (-7.52, -1.56) | 0.003 | 0.53 (-1.48, 2.56) | 0.96 |
| High vs Low Tertile | -4.89 (-8.15, -1.64) | 0.004 | -2.14 (-4.47, 0.20) | 0.07 |
|  | Growth Failure |  |  |  |
|  | OR (95% CI) | P-value | OR (95% CI) | P-value |
| Middle vs Low Tertile | 14.04 (2.94, 66.92) | 0.001 | 0.53 (0.20, 1.38) | 0.20 |
| High vs Low Tertile | 17.22 (3.13, 94.67) | 0.001 | 1.60 (0.57, 4.47) | 0.37 |
| **Sensitivity Analysis #3: Non-Transfer and Survived to S2P (n = 301)** |  |  |  |  |
|  | Daily Weight Gain (gm/day) |  |  |  |
|  | Historical Control |  | ISVMP |  |
|  | Weight Differential (95% CI) | P-value | Weight Differential (95% CI) | P-value |
| Middle vs Low Tertile | -4.13 (-7.09, -1.17) | 0.007 | 0.69 (-1.33, 2.71) | 0.50 |
| High vs Low Tertile | -5.67 (-9.07, -2.28) | 0.001 | -1.96 (-4.29, 0.37) | 0.09 |
|  | Growth Failure |  |  |  |
|  | OR (95% CI) | P-value | OR (95% CI) | P-value |
| Middle vs Low Tertile | 11.34 (2.28, 56.53) | 0.003 | 0.53 (0.20, 1.38) | 0.19 |
| High vs Low Tertile | 18.32 (2.81, 119.33) | 0.002 | 1.59 (0.57, 4.45) | 0.38 |

<sup>a</sup> Controlled for race, ethnicity, maternal age, gender, prematurity, genetic syndrome, birth weight, S1P shunt type, post-S1P ECMO, S1P discharge feeding mechanism, tricuspid regurgitation and right ventricular function prior to S1P discharge, interstage digoxin, and discharge year

